# Searching and visualizing genetic associations of pregnancy traits by using GnuMoM2b

**DOI:** 10.1101/2023.05.25.23290500

**Authors:** Qi Yan, Rafael F. Guerrero, Raiyan R. Khan, Andy A. Surujnarine, Ronald J. Wapner, Matthew W. Hahn, Anita Raja, Ansaf SallebAouissi, William A. Grobman, Hyagriv Simhan, Nathan R. Blue, Robert Silver, Judith H. Chung, Uma M. Reddy, Predrag Radivojac, Itsik Pe’er, David M. Haas

## Abstract

Adverse pregnancy outcomes (APOs) are major risk factors for women’s health during pregnancy and even in the years after pregnancy. Due to the heterogeneity of APOs, only few genetic associations have been identified. In this report, we conducted genome-wide association studies (GWAS) of 479 traits that are possibly related to APOs using a large and racially diverse study, Nulliparous Pregnancy Outcomes Study: Monitoring Mothers-to-Be (nuMoM2b). To display the extensive results, we developed a web-based tool GnuMoM2b (https://gnumom2b.cumcobgyn.org/) for searching, visualizing, and sharing results from GWAS of 479 pregnancy traits as well as phenome-wide association studies (PheWAS) of more than 17 million single nucleotide polymorphisms (SNPs). The genetic results from three ancestries (Europeans, Africans, and Admixed Americans) and meta-analyses are populated in GnuMoM2b. In conclusion, GnuMoM2b is a valuable resource for extraction of pregnancy-related genetic results and shows the potential to facilitate meaningful discoveries.

## Introduction

Pregnancy is a unique window into women’s future health. Adverse pregnancy outcomes (APOs) are frequent and are related to adverse health, such as hypertension, even years after pregnancy (Jowell *et al*. 2022). The burden of APOs varies across racial groups in the United States. For example, Black and Hispanic women have a greater prevalence of APOs than White women (Cho *et al*. 2020). To identify genetic determinants of APOs, we recently conducted genome-wide association studies (GWAS) of four APOs (Guerrero *et al*. 2022) (preterm birth, preeclampsia, gestational diabetes, and pregnancy loss) using samples from a large and racially diverse study, Nulliparous Pregnancy Outcomes Study: Monitoring Mothers-to-Be (nuMoM2b) (Haas *et al*. 2015), gathered from eight clinical centers in the United States. However, few single nucleotide polymorphisms (SNPs) were identified, probably due to the heterogeneity of APOs. For example, preeclampsia is an heterogenous APO characterized by high blood pressure and proteinuria. It can be classified by severity (mild, severe, superimposed, and eclampsia) and onset time (early or late), and dichotomizing it into cases and controls may affect statistical power, as the genetic influence on mild preeclampsia could be closer to controls than other cases. Thus, to provide additional insights into genetic influence on APOs, we conducted GWAS of 479 traits that are possibly related to APOs using nuMoM2b.

To display such extensive results, we have developed a web application, GnuMoM2b (https://gnumom2b.cumcobgyn.org/), for interactively searching, visualizing, and sharing GWAS results. In addition, GnuMoM2b searches for traits associated with any particular SNP to expose pleiotropy across 479 traits (i.e., phenome-wide association study [PheWAS]).

## Results

The GWAS summary statistics of nuMoM2b traits from three individual ancestry group analyses and meta-analyses are populated in GnuMoM2b. The starting page includes usage guidelines. Descriptions of all nuMoM2b traits are provided and searchable by key words. The web interface offers two interactive visualizations: GWAS results queried by trait and PheWAS results queried by SNP. Multiple filters can be used to customize the output, including ancestry selection, *P*-value cutoff, minor allele frequency (MAF) cutoff, and number of studies contributing to meta-analysis. The results can be downloaded in text files. GnuMoM2b also links to NCBI dbSNP to provide additional information on a particular SNP. Furthermore, the genomic control (GC) value will appear after loading a GWAS. The density plot (Figure S1) of GC values from all GWAS indicates well-controlled continuous traits across ancestry-specific and meta-analyses and binary traits in the European (EUR) analysis, with slight deflation in some binary traits in the African (AFR), Admixed American (AMR), and meta-analyses, likely due to smaller sample sizes in AFR and AMR. Therefore, users should consistently check GC values.

GnuMoM2b can facilitate meaningful discoveries. In the online example, GWAS shows that rs988551 in *LAMA2* is the most significant SNP (*P*-value=4.7×10^−9^) associated with gestational hypertension (Trait ID: *acog_PEgHTN_7*) from the meta-analysis (*N*_EUR_=5,636, *N*_AFR_=1,278, and *N*_AMR_=759) (Figure S2), and PheWAS shows that rs988551 is also associated with other hypertension-related traits (*P*-value<1×10^−3^). Lately, *LAMA2* has been implicated to be a preeclampsia-dysregulated gene (Zhou *et al*. 2019). In a recent large GWAS of preeclampsia and gestational hypertension (Honigberg *et al*. 2022), rs167479 in *RGL3* was found to be associated with both conditions in the discovery analysis. Our GnuMoM2b replicates this SNP association at a nominal level (*P*-value=1.2×10^−3^) with gestational hypertension. Note that nuMoM2b participated as a follow-up cohort in that study, but was not included in the discovery analysis. As another example, a large GWAS reported that *WNT4* was associated with gestational length, and the subsequent functional analysis suggested that *WNT4* was a key regulator of decidualization of the human endometrial stromal cell and subsequent embryo implantation (Zhang *et al*. 2017). The rs12037376 in *WNT4* was reported in their paper. As their paper focused on European ancestry, we examined the EUR results in GnuMoM2b. In our PheWAS results, we replicated this SNP at a nominal level (*P*-value=6.8×10^−4^) in association with gestational length (Trait ID: *GAwksEND*). Given that cervical length is a crucial determinant of gestational length (Berghella *et al*. 2003), we further examined the association between this SNP and cervical length at 22-29 weeks (Trait ID: *U3BB02*), resulting in a genome-wide significant *P*-value=1.0×10^−15^ in EUR. In GnuMoM2b, the meta-analysis of GWAS of cervical length reveals that the most significant SNP is rs12404660 (*P*-value=2.3×10^−12^) in *WNT4* (Figure S3), and PheWAS shows its association with gestational length is *P*-value=0.01. However, the association of both rs12037376 and rs12404660 in *WNT4* with gestational length is not significant at the genome-wide level, probably because stillbirth, fetal demise, elective termination, and indicated termination subjects were not excluded from the gestational length analysis. Thus, GnuMoM2b should be used as an exploration tool to search for preliminary results for further analyses. To reach any solid conclusions, refinement of phenotype definitions and statistical analyses as well as experimental validations are needed. Therefore, caution is needed when interpreting the results from GnuMoM2b. Moreover, the nuMoM2b study measured 9 placental analytes at 2 visits (6-13 weeks and 16-21 weeks) searchable under the “Trait Description” tab. The sample sizes (*N*_EUR_>1,000, *N*_AFR_<400, and *N*_AMR_<200) indicate that it could be only reasonable to conduct GWAS in EUR. The GWAS results in EUR show multiple significant loci associated with placental analytes, such as endoglin and sFlt-1 at visit 1, ADAM-12 at visit 2, and fbHCG at both visits (Figure S4). With the advent of summarized GWAS data and two-sample Mendelian randomization (MR) methods, conducting MR analyses have become easier for data scientists. Coupled with external large GWAS of APOs, GnuMoM2b results offer an opportunity to investigate the causal relations between placental analytes and APOs through MR with SNPs as instrumental variables. However, caution is needed to conduct a credible MR analysis and provide a reasonable interpretation.

In addition to searching and visualizing results online, GnuMoM2b allows users to download the summary statistics of three individual ancestry analyses and meta-analyses, which offers users an opportunity to perform summary data-based analysis, such as fine-mapping, colocalization, and MR.

## Discussion

We have conducted GWAS of 479 pregnancy-related traits using the nuMoM2b study and hosted the results on GnuMoM2b that is an easy-to-use web-based application for searching, visualizing, and sharing both genome-wide and phenome-wide association results. The nuMoM2b is a comprehensive cohort with racially diverse participants, offering in-depth clinical and psychosocial phenotyping and longitudinal follow-up during pregnancy. This cohort presents a unique opportunity for a holistic approach to investigating genetic and environmental factors contributing to population morbidity originating in pregnancy. Our GnuMoM2b results complement other large-scale biobank studies, such as the UK Biobank (Bycroft *et al*. 2018), which lack detailed pregnancy phenotypes. These results greatly expand our understanding of genetic influence on pregnancy traits. GnuMoM2b offers researchers, and obstetricians in particular, a new resource to readily extract pregnancy-related genetic results. In the future, we will further add nuMoM2b Heart Health Study (nuMoM2b-HHS) genetic results to GnuMoM2b. The nuMoM2b-HHS, a follow-up study of nuMoM2b, is conducted in a subset of nuMoM2b women, 2-7 years after delivery to better understand the impact of pregnancy outcomes on future health (Haas *et al*. 2016). Lastly, we want to emphasize that GnuMoM2b is a valuable resource for pregnancy data exploration, but understanding the existing biases (e.g., small sample size in particular ancestries, suboptimal trait definitions, and spurious results due to small MAF) is important when interpreting the results, and further analyses are needed to reach any conclusions.

## Materials and methods

The nuMoM2b study enrolled 10,038 women between 2010-2013 from eight centers in the United States. The study aimed to recruit a large and racially diverse cohort of nulliparous pregnant women (Table S1). Its main objective was to identify maternal characteristics, such as genetic factors, physiological responses, and environmental factors that can predict APOs (Haas *et al*. 2015). Participants were followed longitudinally and underwent four study visits from the first trimester to delivery. Throughout pregnancy, various data were collected, such as interviews, questionnaires, research ultrasounds, maternal biometric measurements, and biospecimens (Table S2). The nuMoM2b study methods have been described in detail elsewhere (Haas *et al*. 2015), and the study was approved by the Institutional Review Boards at all participating centers (Guerrero *et al*. 2022). Genome-wide genotyping was conducted using the Infinium Multi-Ethnic Global D2 BeadChip (Illumina, Miami, USA). Quality control measures included assessing sex inconsistencies, autosome missingness, and contamination. Using KING-Robust (Manichaikul *et al*. 2010), kinship was inferred and one participant from each pair with first- or second-degree relatedness was randomly removed. Instead of directly using self-reported race, ancestry was determined using SNPweights, leveraging data from 1000 Genomes Project (Genomes Project *et al*. 2015), resulting in five ancestry groups: EUR (n=6,082), AFR (n=1,425), AMR (n=846), East Asian (EAS; n=323), and South Asian (SAS; n=112). Due to insufficient sample sizes, EAS and SAS were excluded from downstream GWAS. Furthermore, genotype imputation was performed with the TOPMed Imputation Server (https://imputation.biodatacatalyst.nhlbi.nih.gov/). Other details about genome-wide genotyping, genotype imputation, quality control, and ancestry estimation were previously described (Guerrero *et al*. 2022). In this study, after quality control, 8,076 independent subjects (estimated ancestries: 5,891 EUR, 1,374 AFR, and 811 AMR) and 17,177,813 genotyped and imputed SNPs were included in the analyses. The mean age (±standard deviation) at visit 1 was 28.06±5.23 for EUR, 23.42±5.38 for AFR, and 24.75±5.66 for AMR participants. In selecting traits, our aim was to include as many genetically related traits as possible. We first excluded traits that had no genetic relevance, and further filtering was done based on sample size (Figure 1). In the end, we retained 479 traits. Detailed procedures are provided in the supplementary material. The analyses were conducted with PLINK software, version 2 (Chang *et al*. 2015), using linear regression for continuous or ordered categorical traits and logistic regression for binary traits under an additive genetic model, adjusting for maternal age, distance to median maternal age, and the first 10 principal components calculated using genotypic data. For each trait, GWAS was performed in three ancestry groups separately, and only SNPs with MAF>0.01 and Hardy-Weinberg equilibrium (HWE) *P*-value>1×10^−3^ were kept. Next, results from three individual ancestry groups for each trait were pooled using the DerSimonian and Laird method for meta-analyses with random effects implemented in METAL (Willer *et al*. 2010; Hemani 2022), which allows different true effect sizes across three ancestry groups for each SNP.

**Figure 1.**
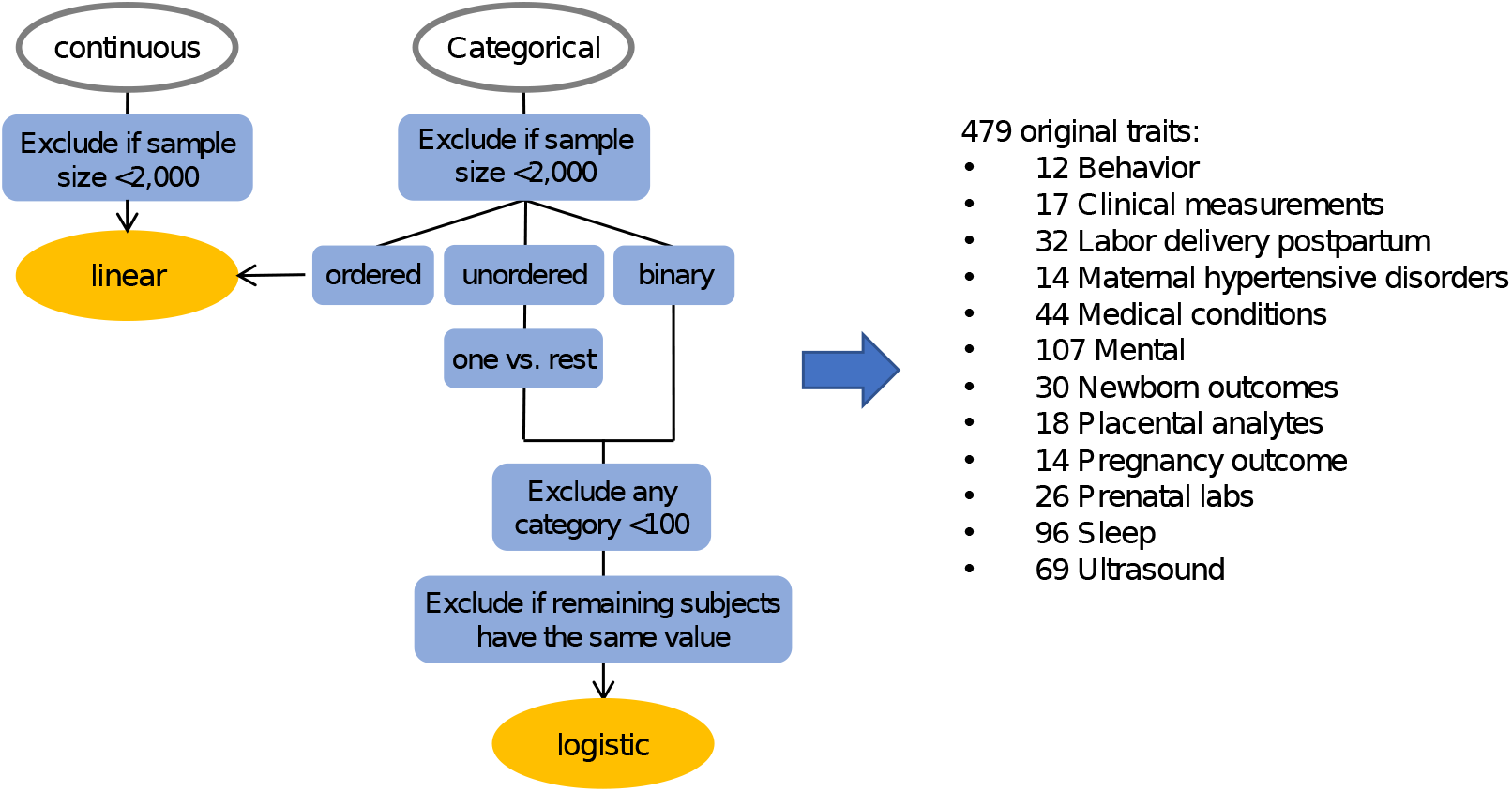
Preprocessing of GWAS traits. The continuous or ordered categorical traits were analyzed using linear regression models. The unordered categorical traits were recoded into several binary traits following the one vs. rest manner, and the binary traits were analyzed using logistic regression models.

Manhattan plots are standard tools to visualize *P*-values of GWAS on a genome-wide level and identify genetic loci associated with the trait. GnuMoM2b is developed to highlight top genetic hits and draw Manhattan plots of GWAS as well as PheWAS using R shiny (http://shiny.rstudio.com/). This web application is easy to use through a graphical user interface.

## Data Availability

The summary-level GWAS and PheWAS data are available and can be downloaded at GnuMoM2b (https://gnumom2b.cumcobgyn.org/). The individual-level phenotype and genotype data have been reported earlier and are available upon request.

## Data availability

The summary-level GWAS and PheWAS data are available and can be downloaded at GnuMoM2b (https://gnumom2b.cumcobgyn.org/). The individual-level phenotype and genotype data have been reported earlier (Haas *et al*. 2015; Guerrero *et al*. 2022) and are available upon request.

## Funding

Support for performing DNA extraction and GWAS from the Indiana University Grand Challenges Precision Diabetes project funding. D.M.H. was partially funded by R01HD101246 from the U.S. National Institutes of Health (NIH). Original funding for nuMoM2b sample and data collection are noted in the study methods paper (Haas *et al*. 2015).

## Conflict of interest

None declared.

## Notes

### Competing Interest Statement

The authors have declared no competing interest.

### Author Declarations

Nulliparous women with singleton pregnancies were recruited from hospitals affiliated with eight clinical centers: Case Western University; Columbia University; Indiana University; University of Pittsburgh; Northwestern University; University of California at Irvine; University of Pennsylvania; and University of Utah. The Data Coordinating and Analysis Center is RTI International. Each site's local governing Institutional Review Board(s) approved the protocol and procedures.

