## Supplementary material for "Searching and visualizing genetic associations of pregnancy traits by using GnuMoM2b"

**Trait selection**

Our aim was to include as many genetically related traits as possible, excluding trisomy and race-related variables. We started with all variables, then first excluded administrative ones, such as IDs, dates, sites, consent, and eligibility. We also excluded all medication variables, traits for family members (not for the pregnant participants), socioeconomic variables, etc. The goal was to retain only genetically related traits, acknowledging that some traits are likely more genetically relevant than others. Next, the traits were further filtered out based on sample size, as shown in Figure 1. We excluded traits if the non-missing sample size was less than 2000. We transformed each unordered categorical trait into several binary traits following the one-vs-rest manner. Any category in a binary trait with fewer than 100 samples was removed. If all remaining values belonged to the same category, we then excluded the trait. This process resulted in a final set of 479 traits.

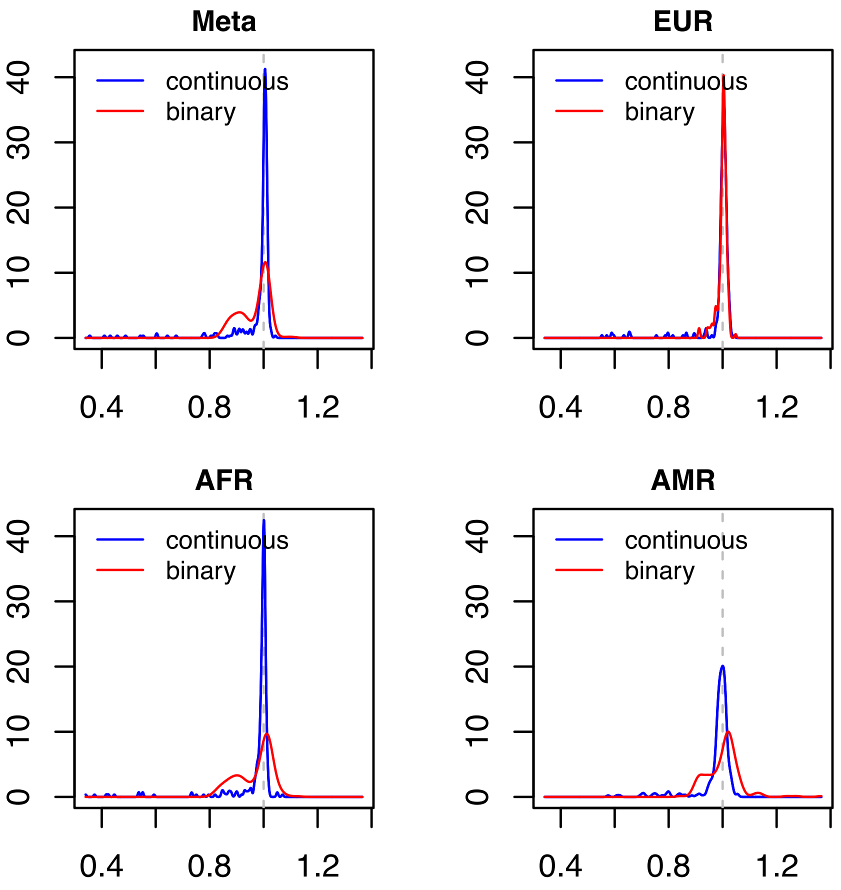


**Figure S1. Density plots of genomic control values from all GWAS.** The blue curve represents GWAS of continuous traits, and the red curve represents GWAS of binary traits. The x-axis displays the genomic control values, and the y-axis displays the corresponding density values. Note that density values are not actual probabilities but probability densities.

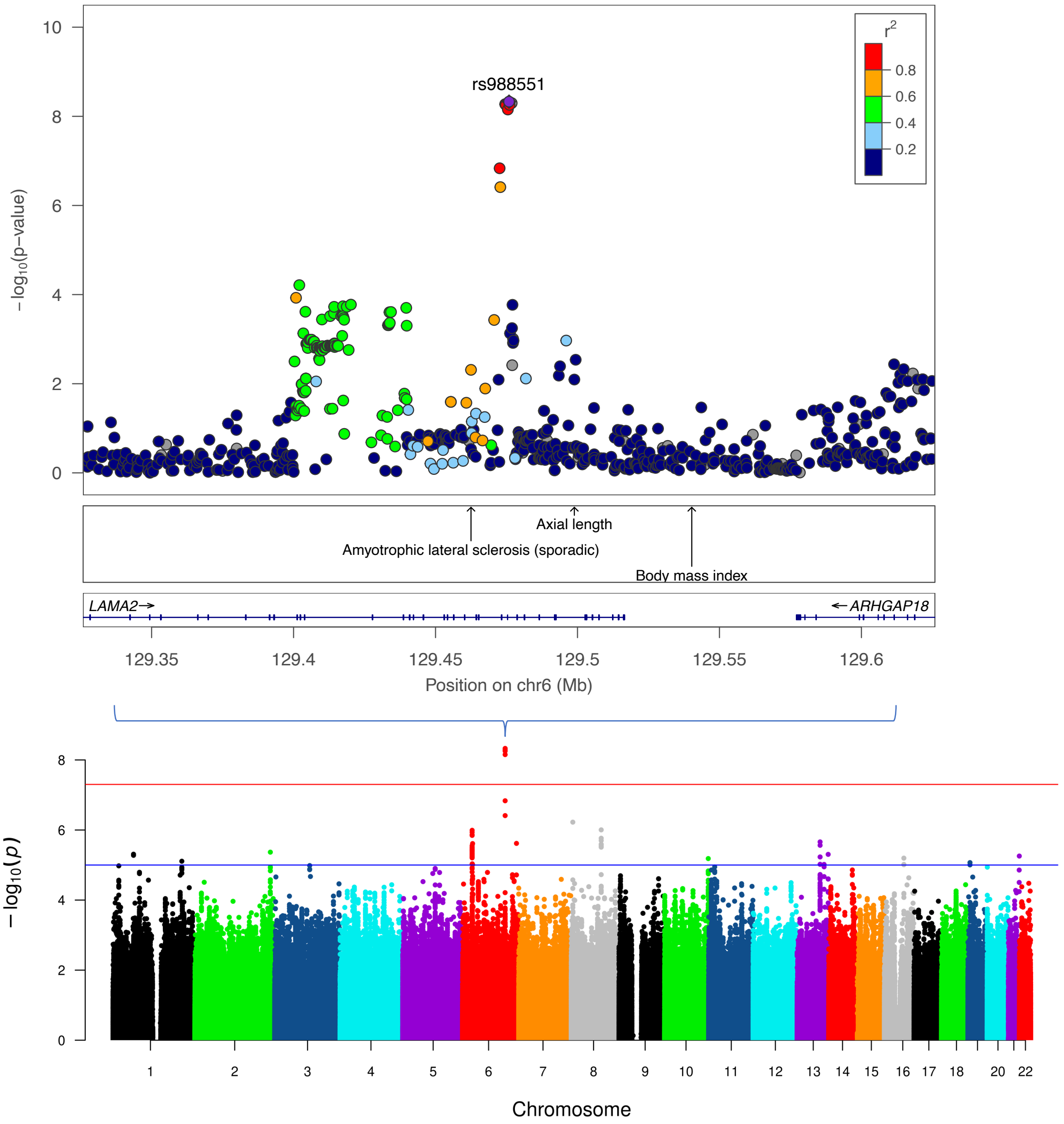


**Figure S2. GWAS of gestational hypertension from the meta-analysis.** The bottom panel is Manhattan plot. The chromosomal position of each SNP is displayed along the x-axis and the negative log10 of the association *P*-value is displayed on the y-axis. The red line represents the genome-wide significance level ($P < 5\times10^{-8}$) and blue line represents the suggestive significance level ($P < 1\times10^{-5}$). The top panel is regional plot for the top SNP (rs988551). The lower portion of the figure displays the relative location of genes and the direction of transcription, while the middle portion shows known GWAS associations at the locus from the GWAS catalog. The x-axis displays the chromosomal position and the y-axis shows the significance of the associations. The purple diamond shows the *P*-value rs988551. The circles show the *P*-values for all other SNPs and are color coded according to the level of LD with rs988551 using combined nuMoM2b EUR, AFR, and AMR individuals. Only SNPs present in EUR, AFR, and AMR are displayed.

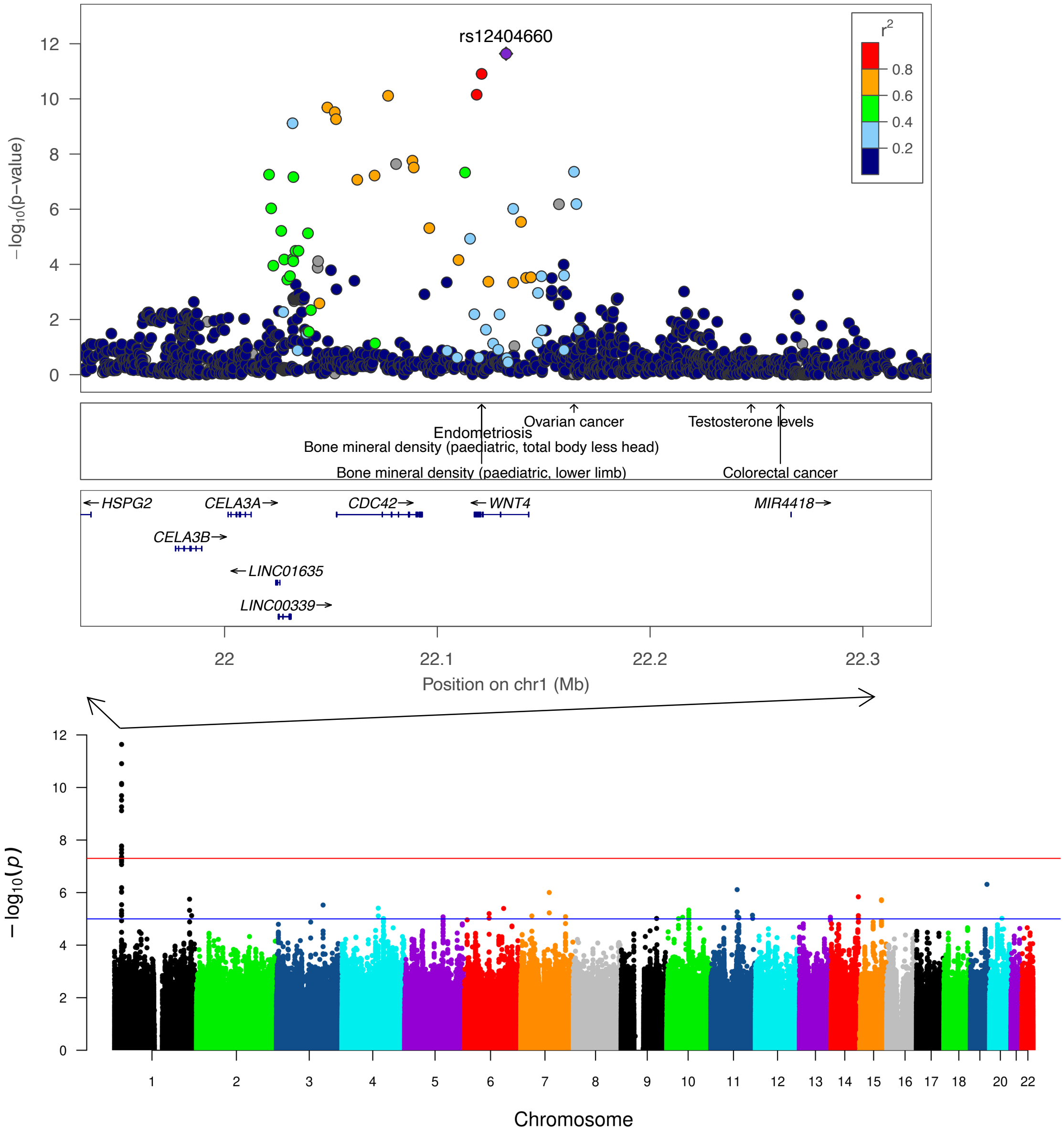


**Figure S3. GWAS of cervical length at 22-29 weeks from the meta-analysis.** The bottom panel is Manhattan plot. The chromosomal position of each SNP is displayed along the x-axis and the negative log10 of the association *P*-value is displayed on the y-axis. The red line represents the genome-wide significance level ($P < 5{\times}10^{-8}$) and blue line represents the suggestive significance level ($P < 1{\times}10^{-5}$). The top panel is regional plot for the top SNP (rs12404660). The lower portion of the figure displays the relative location of genes and the direction of transcription, while the middle portion shows known GWAS associations at the locus from the GWAS catalog. The x-axis displays the chromosomal position and the y-axis shows the significance of the associations. The purple diamond shows the *P*-value for rs12404660. The circles show the *P*-values for all other SNPs and are color coded according to the level of LD with 12404660 using combined nuMoM2b EUR, AFR, and AMR individuals. Only SNPs present in EUR, AFR, and AMR are displayed.

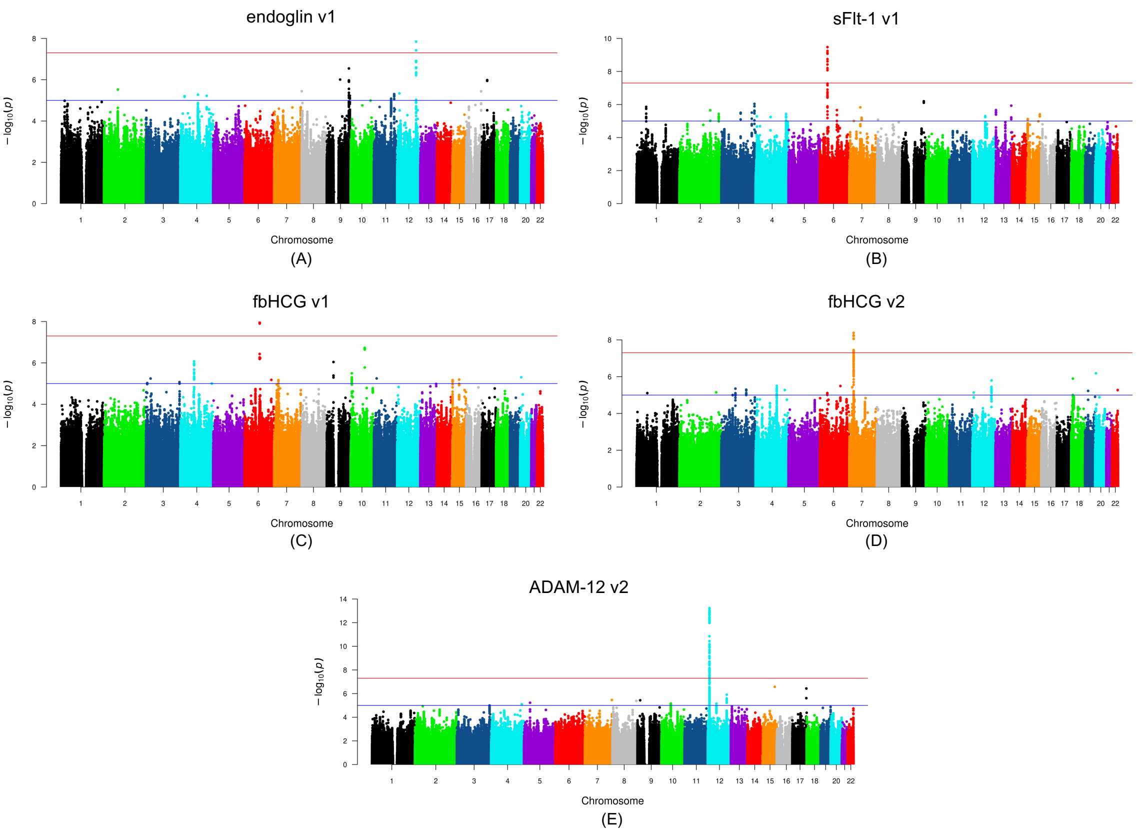


**Figure S4. GWAS of placental analytes using individuals of white ancestry.** Manhattan plots for (A) endoglin (visit 1), (B) sFlt-1 (visit 1), (C) fbHCG (visit 1), (D) fbHCG (visit 2), and (E) ADAM-12 (visit 2). The chromosomal position of each SNP is displayed along the x-axis and the negative log10 of the association *P*-value is displayed on the y-axis. The red line represents the genome-wide significance level ($P < 5.6\times10^{-9}$) and blue line represents the suggestive significance level ($P < 5\times10^{-8}$). Only SNPs with MAF>0.05 are displayed.

Table S1. Self-reported race of nuMoM2b participants

| **Baseline Characteristics** | **nuMoM2b (n=10,038)** |
|---|---|
| *Maternal race n (%)* | |
| White non-Hispanic | 5,989 (59.7%) |
| Black non-Hispanic | 1,418 (14.1%) |
| Hispanic | 1,700 (16.9%) |
| Asian | 407 (4.1%) |
| Other | 524 (5.2%) |

Table S2. Timing of data collection in nuMoM2b

| Question Domains, Samples, and Clinical Evaluations | Pregnancy Trimester (nuMoM2b)* | | | |
|---|---|---|---|---|
| | Visit 1 | Visit 2 | Visit 3 | Delivery |
| Demographic characteristics | X | X | X | X |
| Medical history | X | X | X | X |
| Psychological factors | X | X | X | |
| Biometric measurements | X | X | X | |
| Ultrasound | X | X | X | |
| Biospecimens | | | | |
| Urine | X | X | X | |
| Blood (whole blood, plasma, serum) | X | X | X | X |
| Cervicovaginal fluid | X | X | X | X |
| Cord blood (whole blood, plasma, serum), Neonatal saliva | | | | X |
| Placenta, fetal membranes, umbilical cord segment | | | | X |
| Symptoms or diagnoses between the visit 3 and the admission for delivery | | | | X |
| Participant assessment of delivery route/reasons | | | | X |

* Study visits were during the following gestational age intervals: first trimester, 6 weeks 0 days to 13 weeks 6 days; second trimester, 16 weeks 0 days to 21 weeks 6 days; and third trimester, 22 weeks 0 days to 29 weeks 6 days.
